# Clinical utility of home versus hospital spirometry in fibrotic ILD: evaluation following INJUSTIS interim analysis

**DOI:** 10.1101/2021.05.20.21257328

**Authors:** Fasihul Khan, Lucy Howard, Glenn Hearson, Colin Edwards, Chris Barber, Steve Jones, Andrew M Wilson, Toby M Maher, Gauri Saini, Iain Stewart, Gisli Jenkins

## Abstract

The COVID-19 pandemic identified an urgent need to re-evaluate the provision of spirometry for clinical monitoring. Home spirometry offers the opportunity for real-time disease evaluation without risk of nosocomial infection. To determine the utility of home spirometry in interstitial lung disease (ILD), interim data from the ongoing INJUSTIS study was evaluated. High correlation was observed between home and hospital spirometry at baseline(r=0.89) and three-months(r=0.82). Over 90% of home spirometry values were within Bland-Altman agreement limits at both time points, although frequently underestimated hospital values. Home spirometry is feasible in people with fibrotic ILD.

## Introduction

Domiciliary monitoring of physiological variables has become routine in many chronic conditions owing to technological advances(1). Restricted clinical capacity and patient safety during the COVID-19 pandemic have identified an urgent need to consider remote lung function monitoring of chronic respiratory disease(2). Home handheld spirometry enables repeated measurements, offering opportunities for real-time disease evaluation, without the risk of nosocomial infection.

Interstitial lung disease (ILD) encompasses a heterogeneous range of immuno-inflammatory and fibrotic diseases. Forced vital capacity (FVC) correlates with outcome in ILD and remains the most commonly used biomarker of disease progression(3), with clinical trials consistently adopting hospital FVC measurements as the primary endpoint(4-6). Recent studies using home spirometry support feasibility in idiopathic pulmonary fibrosis (IPF), but little data exists regarding the acceptance of daily spirometry in non-IPF ILD and its comparability to hospital spirometry (7-10).

We assessed interim data from the It’s Not Just Idiopathic Pulmonary Fibrosis Study (INJUSTIS, NCT03670576) (11) to evaluate the clinical utility of home spirometry as an alternative to hospital spirometry in participants with fibrotic ILD.

## Methods

Participants diagnosed with multidisciplinary team confirmed fibrotic ILD (unclassifiable, hypersensitivity pneumonitis, asbestosis, rheumatoid-associated ILD and IPF) were recruited. Participants were offered a portable handheld spirometer (MIR Spirobank Smart) linked via Bluetooth to a smartphone application and asked to perform a single, blinded forced expiratory manoeuvre daily for at least three months. Hospital spirometry was collected according to international guidelines,(12) and was obtained as standard of care at baseline and at a three month research visit. Detailed INJUSTIS protocol information is available(11).

Home spirometry readings falling within the upper and lower centile of aggregated group data based on FVC %predicted values were excluded to limit effects of substandard blows. A mean value based on measures over a week was calculated for comparison to the corresponding hospital value, baseline was defined as day(d)1 to d7 and three months, d99 to d105.

Correlation coefficients between home and hospital spirometry for corresponding timepoints were assessed using Pearson correlation. Bland-Altman plots were generated to assess the number of measurements that were outside the 95% limits of agreement. We assessed overall adherence of daily home spirometry and consistency of measures across each week of study. A weekly coefficient of variation was calculated where three or more daily values were provided, which was assessed in a generalised estimating equation population-averaged model with exchangeable correlation matrix and robust sandwich variance estimators. Association of subgroup, week and interaction of week and subgroup were estimated. All analyses were performed using Stata v.16 (StataCorp, College Station, TX, USA).

## Results

Eighty-two participants were included in analysis, of which 23 had IPF (28%) and 59 had non-IPF ILD (72%). Thirty-five participants had three-month data for both home and hospital spirometry (Table 1). Mean age was 69.8±8 years, 72.3% were male and mean FVC was 2.96±0.88L. Median adherence to daily home spirometry was 81% (IQR 61-94%).

**Table 1.** Comparison of FVC shown in litres after FVC percent predicted <1^st^ and >99^th^ centile excluded. Values shown for all patients, and for non-IPF ILD separately. Agreement after values plotted on Bland-Altman plot, with N the total number of participants with values outside limits. Correlation presented between hospital and home spirometry.

|  |  | Comparison |  |  | Agreement |  | Correlation |  |  |
| --- | --- | --- | --- | --- | --- | --- | --- | --- | --- |
| FVC sample | N | Mean Lab (SD) | Mean Home (SD) | Mean diff (SD) | N Outside limits | % Within limits | r | R2 | P |
| All |  |  |  |  |  |  |  |  |  |
| Baseline | 82 | 2.96 (0.88) | 2.71 (0.88) | -0.25 (0.40) | 7 | 91.5 | 0.89 | 0.79 | <0.0001 |
| 3 months | 35 | 2.96 (0.98) | 2.84 (0.99) | -0.12 (0.59) | 1 | 97.1 | 0.82 | 0.67 | <0.0001 |
| Non-IPF ILD only |  |  |  |  |  |  |  |  |  |
| Baseline | 59 | 2.8 (0.82) | 2.57 (0.85) | -0.23 (0.39) | 4 | 93.2 | 0.89 | 0.79 | <0.0001 |
| 3 months | 26 | 2.83 (0.95) | 2.65 (0.83) | -0.18 (0.57) | 1 | 96.2 | 0.81 | 0.66 | <0.0001 |

Of the total 6202 daily FVC measurements, values in the upper and lower centile (below 27.15% or above 144.17% predicted FVC) were excluded. High correlation was observed between home and hospital spirometry at baseline (r=0.89) and three-months (r=0.82) (Table 1). Bland-Altman plots demonstrated more-than 90% of home spirometry values were within agreement limits of hospital values at both timepoints (Figure 1). Home values more frequently underestimated hospital values. Similar results were obtained when restricted to non-IPF participants specifically.

**Figure 1:**
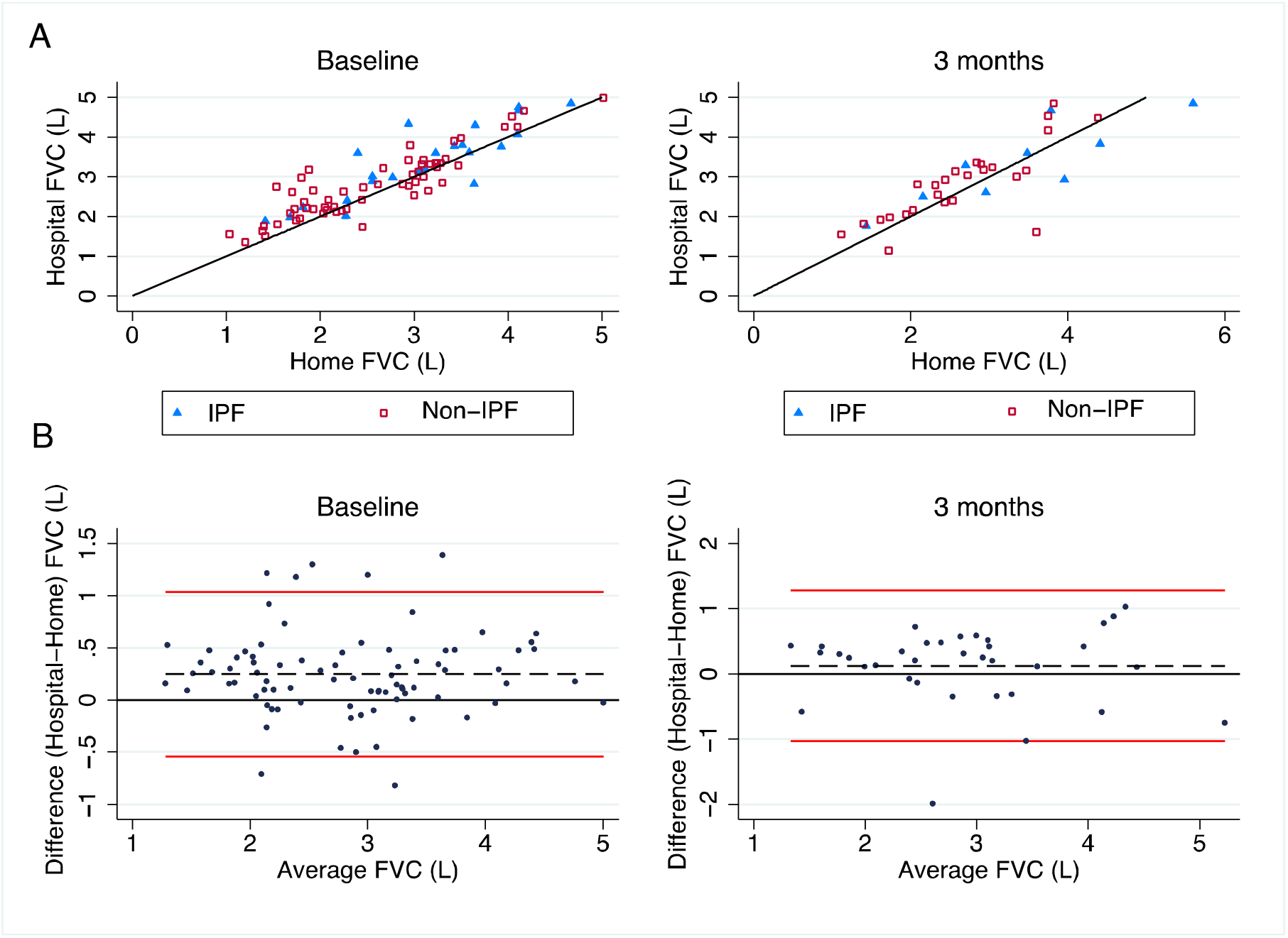
A. Correlation of home and hospital FVC (litres) measurements at baseline and 3 months, coloured differently for IPF (n=23 at baseline; n=9 at 3 months) and non-IPF (n=59 at baseline; n=26 at 3 months). Black reference line represents y=x. B. Bland Altman plot for baseline and 3 months. Mean difference of hospital relative to home spirometry was 0.25L (SD 0.4) at baseline and 0.12L (SD 0.59) at 3 months. The red lines represent the 95% limits of agreement. Baseline measurements were calculated as the mean of daily readings obtained during the first seven days. Three-month measurements were calculated as the mean of readings obtained between days 99 and 105.

A slightly higher coefficient of variation (CoV) was observed in the phenotypically more diverse and larger non-IPF ILD subgroup, although no significant association with CoV was observed in longitudinal analysis (coefficient 2.11, 95%CI -1.60;5.83, p=0.144) (Figure 2). Overall, weekly CoV did not significantly change (−0.22, 95%CI -0.52;0.08, p=0.144), indicating that weekly averages reliably reflect daily values for comparison to a single time point of hospital spirometry. A suggestive, non-significant reduction in variability over time may be attributable to learning and improved technique. No interaction with ILD subgroup was observed at any week.

**Figure 2:**
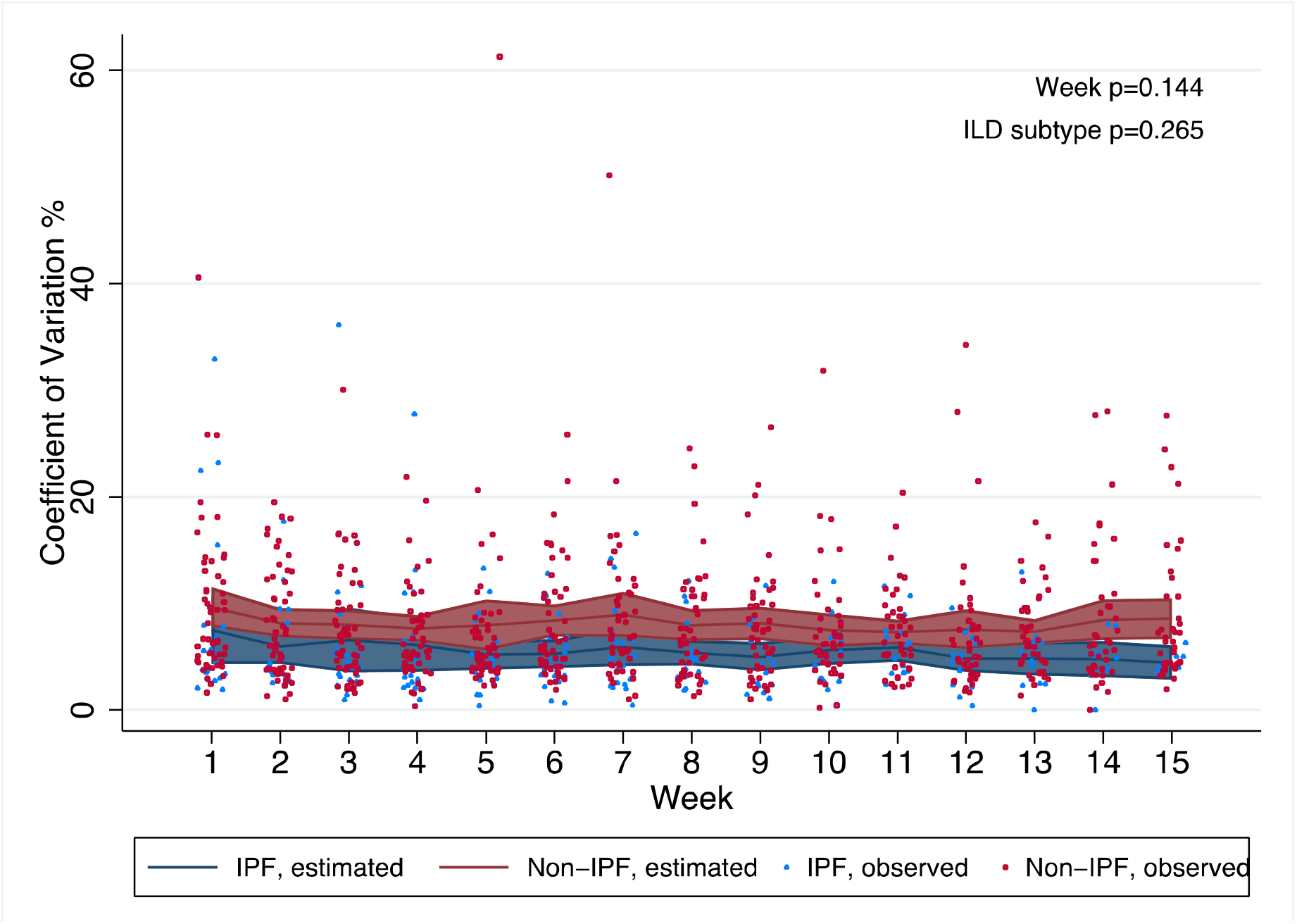
Weekly coefficient of variation (CoV) (%) in home spirometry across study time for ILD subtype. Blue and red lines represent estimated CoV (and 95% confidence intervals) in IPF and non-IPF group, respectively. Scatter points for observed individual participant weekly CoV. Number of participants included at each week (p-value for ILD subtype interaction): week 1, 76 (0.987); week 2, 72 (0.946); week 3, 73 (0.695); week 4, 69 (0.790); week 5, 70 (0.756); week 6, 69 (0.574); week 7, 68 (0.617); week 8, 65 (0.791); week 9, 63 (0.619); week 10, 59 (0.903); week 11, 58 (0.734); week 12, 58 (0.742); week 13, 55 (0.842); week 14, 52 (0.490); week 15, 46 (0.391). P values from generalised estimating equation shown for change in coefficient of variation per week, and ILD subtype (IPF and non-IPF).

## Discussion

Our findings support the clinical utility of home spirometry in the remote monitoring of patients with ILD. Although participants were blinded, adherence to daily spirometry remained high, and was similar to adherence rates in non-blinded studies(13). We stipulated the performance of daily measures rather than a minimum number of weekly blows,(8, 9) with reliable adherence in the three-month design. Home and hospital measurements were highly correlated at complementary time points, though home spirometry tended to underestimate measurements when compared with hospital spirometry(7). The mean difference at baseline was 0.25L lower with over 90% of measurements within agreement limits. Limitations of using home measures as a direct surrogate for hospital spirometry are highlighted where agreement limits were not met. Furthermore, although variability was observed, daily measures indicated minimal influence of time or disease. Whilst we demonstrate comparability of measurements, we emphasise the importance of longitudinal modelling of daily spirometry for clinical endpoint precision.

Our study was limited by modest interim sample sizes and a restricted follow up due to interim censoring attributable to the COVID-19 pandemic. Participants were asked to perform a single reading without replication to minimise potential intrusiveness of multiple daily expiratory manoeuvres. Baseline hospital spirometry was obtained pragmatically as a standard of care, the acceptable timeframe from recruitment may have contributed to larger discrepancies with home spirometry at this time point compared with three-month research visits. We were unable to validate the quality of participant attempts as the handheld device did not record flow-volume loops. It is likely these factors would be compensated in longitudinal modelling of daily spirometry, whilst the intention here was to assess comparability to hospital spirometry when evaluated as a single value.

In summary, we demonstrate that blinded, daily home spirometry in fibrotic ILD irrespective of aetiology or subtype, is feasible, reliable and within acceptable levels of agreement to hospital spirometry for clinical measurement. This is likely to be particularly relevant where clinical access is limited due to geographical factors, patient choice, service pressures and future pandemics.

## Data Availability

Ongoing study

